# COVID-19 convalescent plasma for the treatment of immunocompromised patients: a systematic review

**DOI:** 10.1101/2022.08.03.22278359

**Authors:** Jonathon W. Senefeld, Massimo Franchini, Carlo Mengoli, Mario Cruciani, Matteo Zani, Ellen K. Gorman, Daniele Focosi, Arturo Casadevall, Michael J Joyner

## Abstract

Immunosuppressed patients have increased risk for morbidity and mortality from COVID-19 because they less frequently mount antibody responses to vaccines and often cannot tolerate small-molecule antivirals. The Omicron variant of concern of SARS-CoV-2 has progressively defeated anti-Spike mAbs authorized so far, paving the way to a return to COVID-19 convalescent plasma (CCP) therapy. In this systematic review we performed a metanalysis of 9 controlled studies (totaling 535 treated patients and 1365 controls and including 4 randomized controlled trials), an individual patient data analysis of 125 case reports/series (totaling 265 patients), and a descriptive analysis of 13 uncontrolled large case series without individual patient data available (totaling 358 patients). The metanalysis of controlled studies showed a risk ratio for mortality of 0.65 (risk difference -0.11) in treatment with CCP versus standard of care for immunosuppressed COVID-19 patients. On the basis of this evidence, we encourage initiation of high-titer CCP from vaccinees(‘hybrid plasma’) in immunocompromised patients.

## Introduction

In December 2019, severe acute respiratory syndrome coronavirus-2 (SARS-CoV-2) emerged in Wuhan (China)^1,2^, causing coronavirus disease 2019 (COVID-19). It rapidly spread across the globe leading to a pandemic with currently nearly 577 million infected people worldwide and 6.4 million deaths ^3^. A number of treatments, including antiviral, anticoagulant and anti-inflammatory agents have been tested in COVID-19 patients, with often controversial results ^4^. The passive transfer of anti-SARS-CoV-2 neutralizing antibodies (nAb) from plasma of recently recovered individuals (COVID-19 convalescent plasma, CCP) to patients with severe COVID-19 was among the first therapies used ^5-7^. Nowadays we know that such antibody-based treatment, when administered early (within 72 hours since onset of symptoms) and with high titers of neutralizing antibodies (nAb), leads to clinical benefit ^8,9^.

In spite of the marketing of monoclonal antibodies (mAbs) against COVID-19 since February 2021, the high mutation rate of SARS-CoV-2 (approximately a novel variant of concern (VOC) every 6 months) progressively escaped most, if not all, mAbs ^10^. By contrast, thanks to the prompt availability of CCP against emerging VOCs, CCP maintained its efficacy over time by following the ongoing Spike protein evolution. As a result, since the beginning of 2022 there has been a renewed interest toward CCP use, particularly for immunocompromised patients, who are not able to mount a sufficiently protective antibody response against the virus and have contraindications or side effects from small molecule antivirals ^11,12^. These patients are at higher risk for morbidity and mortality from COVID-19 ^13^. A few controlled studies and a number of case reports and case series have shown a clinical benefit from CCP use for COVID-19 patients with hematological or solid cancer or other underlying causes of immunosuppression. For such reasons, on January 2022 the US Food and Drug Administration (FDA) revised the Emergency Use Authorization (EUA) of CCP to include those hospitalized with impaired humoral immunity ^14^.

To further summarize the heterogeneous literature data on the CCP use in immunocompromised patients, possibly identifying factors involved in more favorable outcomes, we have performed here a systematic literature review.

## Materials and methods

### Search criteria

In this systematic review, we investigated the impact of CCP on COVID-19 mortality in patients with primary (i.e., inheritable) or secondary immunosuppression (i.e., related to haematological or solid cancers, autoimmune disorders or organ transplants). For this purpose, an electronic literature search through the online PubMed and MEDLINE databases was initiated for articles published from January 1, 2020 to August 12, 2022, using English language as the only restriction. The Medical Subject Heading (MeSH) and keywords used were: (“COVID-19” OR “SARS-CoV-2” OR “coronavirus disease 2019”) AND (“convalescent plasma” OR “immune plasma” OR “hyperimmune plasma”) AND (“immunosuppression” OR “immunodeficiency” OR “immunocompromised” OR “cancer” OR “transplant” OR “malignancy” OR “hematological” OR “oncologic” OR “lymphoma” OR “leukemia” OR “myeloma” OR “agammaglobulinemia” OR “hypogammaglobulinemia” OR “common variable immunodeficiency” OR “autoimmune disorder”). Relevant articles and data were also identified through non-systematic searches in Google Scholar and medRxiv, including abstracts of congress presentations which were not published yet. We also screened the reference list of reviewed articles for additional studies not captured in our initial systematic literature search. To be considered eligible for inclusion, studies must include: 1) patients with primary or secondary immunosuppression with a confirmed diagnosis of COVID-19; 2) CCP as a COVID-19 treatment; and 3) information on patients’ outcome. To perform a comprehensive analysis, the retrieved literature was grouped into three different strata, according to information characteristics: 1) controlled trials underwent a quantitative analysis (meta-analysis); 2) large case series with aggregated data underwent a descriptive analysis; and 3) case reports and case series with individual patient data underwent a single patient analysis.

Articles underwent a reciprocally blind evaluation for inclusion by two assessors (J.W.S. and M.F.) and disagreements were resolved by a third senior assessor (D.F.). A PRISMA flowchart for this review is available in Figure 1. The following data, when available, were searched for each case: patient’s sex and age, the underlying primary or secondary immunodeficiency, the 11-point WHO COVID-19 disease severity score ^15^, the need for mechanical ventilation, survival at the end of follow-up, the number of CCP units transfused, the volume of each CCP unit, the total CCP volume transfused, the antibody level (either nAb titer or anti-Spike IgG levels) and the antibody test used, time from admission to CCP transfusion, time from symptom onset to CCP transfusion, rapid clinical improvement (defined as a reduction in supplemental oxygen requirements < 5 days post-CCP transfusion), duration of follow-up (days), need for admission to intensive care unit (ICU), ICU length of stay (days; total and after CCP transfusion), concomitant COVID-19 antiviral treatments (intravenous immunoglobulins, remdesivir, hydroxychloroquine, anti-Spike mAbs), and specific immunosuppressive drugs (anti-CD20 mAbs).

**Figure 1.**
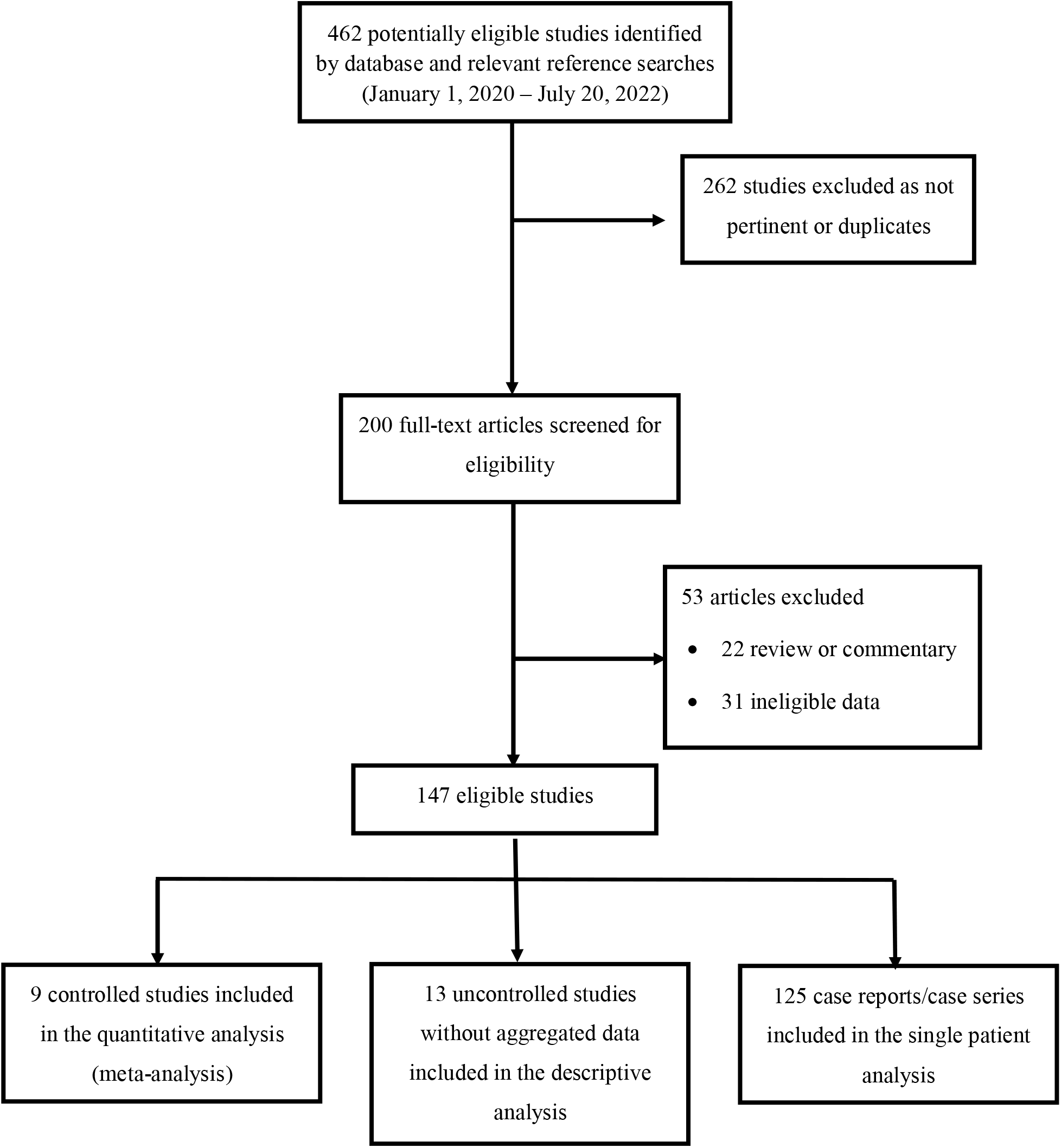
PRISMA flow chart for the current study.

### Systematic review of comparative studies

We have considered both RCTs and non-RCT. Two review authors (MF, MC) independently assessed the risk of bias (ROB) of each included study following the domain-based evaluation described in the *Cochrane Handbook for Systematic Reviews of Interventions*. [Higgins, J.P., Green, S. (eds.): Cochrane Handbook for Systematic Reviews of Interventions – Version 5.1.0 [updated March 2011]. The Cochrane Collaboration http://www.cochranehandbook.org.].

Within-trial ROB was assessed, using the Cochrane ROB tool for RCTs and the ROBINS-I tool for non-RCTs.[Higgins, J.P., Green, S. (eds.): Cochrane Handbook for Systematic Reviews of Interventions – Version 5.1.0 [updated March 2011]. The Cochrane Collaboration http://www.cochranehandbook.org.]. The Cochrane ‘Risk of bias’ tool for RCTs addresses six specific domains: sequence generation, allocation concealment, blinding, incomplete data, selective outcome reporting, and other issues relating to bias. The methodological quality of observational studies was assessed with the ROBINS-1 tool ^16^. This tool includes seven specific bias domains, pre-intervention and post-intervention. The domains are: (1) confounding; (2) selection of participants; (3) classification of intervention; (4) deviation from interventions (o biases that arise when there are systematic differences between the care provided to experimental intervention and comparator groups, beyond the assigned interventions); (5) missing outcome; (6) measurement of outcomes; (7) selection of reported result overall. For both RCTs and non-RCTs we have presented our assessment of risk of bias using two ‘Risk of bias’ summary figures: 1) a summary of bias for each item across all studies; and 2) a cross-tabulation of each trial by all the ‘Risk of bias’ items.

### Effect of intervention

Measures of treatment effect were risk ratio (RR) and mean difference (MD). The study weight was calculated using the Mantel-Haenszel method. We assessed statistical heterogeneity using t^2^, Cochran’s Q and *I*^*2*^ statistics ^17^. The *I*^*2*^ statistic describes the percentage of total variation across trials due to heterogeneity rather than sampling error. In the case of no heterogeneity or moderate heterogeneity (*I*^2^<40), studies were pooled using a fixed-effects model. Where values of *I*^2^ were >40, a random-effects analysis was undertaken.

### ’Summary of findings’ tables

For the outcome mortality, we used the principles of the GRADE system to assess the quality of the body of evidence associated with specific outcomes and constructed ‘Summary of findings’ tables using REVMAN 5.4 ^18,19^. These tables present key information concerning the certainty of the evidence, the magnitude of the effects of the interventions examined, and the sum of available data for the main outcomes. The ‘Summary of findings’ tables also include an overall grading of the evidence related to each of the main outcomes using the GRADE approach, which defines the certainty of a body of evidence as the extent to which one can be confident that an estimate of effect or association is close to the true quantity of specific interest. The certainty of a body of evidence involves consideration of within-trial risk of bias (methodological quality), directness of evidence, heterogeneity, precision of effect estimates, and risk of publication bias.

### Statistical analysis of individual patient data

In the descriptive statistics of individual patient data, continuous variables were reported as mean (+SD) or median (range) as appropriate according to distribution, while categorical variables were reported as numbers and percentages.

An exploratory analysis on the main theme was done on the mortality after discretizing the total volume, as <= 200 ml to 1800 ml with serial increases of 200 ml. Then, a breakpoint at 600 ml of CCP was tentatively posed, allowing to compare the mortality when the CCP was under or over the level by Fisher exact test.

The basic model consisted in a logistic regression using mortality as dependent variable, and total volume as predictor. The CCP total volume was expressed in units of 100 ml (“totvol100”), for ease of interpretation. The potential additive independent effects of “Age”, “Sex”, “Time from admission to transfusion”, “Rapid improvement (within 5 days)”, “ICU length of stay”, “Steroids”, “Remdesivir”, “(Hydroxy)chloroquine”, “Antibiotics”, “Anti-CD20”, and “Category” were evaluated.

In power analysis, the total sample size was calculated in order to detect an experimental-group proportion of 0.06 as death rate, with the control-group proportion of 0.08, assuming a one-sided hypothesis test with a 5% significance level, focusing a desired power of 80%, and if both groups (treated and untreated) had the same number of observations. This would correspond to the prevention of 25% of the basal deaths, or a risk ratio (RR) of 0.75. Stata 17.0 was used for all statistical calculations.

## Results

The literature search yielded 147 eligible studies, of which 9 controlled trials (4 randomized trials ^20-23^ and 5 cohort studies ^24-28^) were selected for meta-analysis, 13 uncontrolled large case series without individual patient data but totaling 358 patients were selected for descriptive analysis ^29-41^, and 125 case reports/series totaling 265 patients met the eligibility criteria for individual patient data analysis, ^35,42-165^. Reference ^35^ was included in both the descriptive analysis and the individual patient data analysis because individual patient data were available only for a subgroup of patients.

In the metanalysis of the 9 controlled trials (totaling 535 patients treated with CCP and 1365 controls), the main findings are summarized in Table 1. As shown in Figure 2 the level of evidences was generally moderate because of a moderate ROB (Figure 2). Despite this, there is a high level of concordance among study outcomes, with a risk difference of -0.11, and a risk ratio of 0.65 (Figure 3). We note that several cohort studies, despite being defined as propensity-score matched^28^, did not match for concomitant antivirals (potentially masking CCP efficacy) or B-cell depleting agents. In sensitivity analysis, exclusion of individual studies did not affect significantly the effect size.

**Table 1.** Summary of findings for the 9 controlled studies included in the metanalysis.

| Patient or population: Immunocompromised patients |  |  |  |  |  |  |
| --- | --- | --- | --- | --- | --- | --- |
| Settings: Hospitalized patients with COVID-19 |  |  |  |  |  |  |
| Intervention: COVID-19 convalescent plasma (CCP) |  |  |  |  |  |  |
| Comparison: standard of care (SOC) |  |  |  |  |  |  |
| Outcomes | Illustrative comparative risks* (95% CI) |  | Relative effect (95% CI) | No of Participants (studies) | Quality of the evidence (GRADE) | Comments |
|  | Assumed risk | Corresponding risk |  |  |  |  |
|  | Controls (standard of care) | Intervention (convalescent plasma) |  |  |  |  |
| - All studies (RCTs and non-RCTs) | 281 per 1000 | 183 per 1000 (from 152 to 222) | RR 0.65 (95% CIs, 0.54 to 0.79) | 1900 patients (9 trials, 4 RCTs and 5 non-RCTs) | ⊕⊕⊖⊖ low (downgraded twice for serious ROB) | Mortality was observed more commonly among SOC recipients compared to CP |
| -RCTs only | 281 per 1000 | 183 per 1000 (from 152 to 222) | RR 0.68 (95 % CIs, 0.51/0.91) | 340 participants (4 RCTs) | ⊕⊕⊕⊖ moderate (downgraded for ROB) | CP reduces mortality compared to SOC |
| -Cohort studies only | 264 per 1000 | 169 per 1000 (from 132 to 216) | RR 0.64 (95 % CIs, 0.50/0.82) | 1560 participants (5 trials) | ⊕⊕⊖⊖ low (downgraded twice for serious risk of bias) | Mortality was observed more commonly among SOC recipients compared to CCP. In sensitivity analysis, exclusion of individual studies did not affect the effect size of intervention. |
| - All studies | 281 per | 183 per 1000 | RR 0.65 | 1900 | ⊕⊕⊖⊖ | Mortality was |
| (RCTs and non-RCTs) | 1000 | (from 152 to 222) | (95% CIs, 0.54 to 0.79) | patients (9 trials, 4 RCTs and 5 non-RCTs) | low (downgraded twice for serious ROB) | observed more commonly among SOC recipients compared to CCP |

\*The basis for the assumed risk is the mean control group risk across studies. The corresponding risk (and its 95% confidence interval) is based on the assumed risk in the comparison group and the relative effect of the intervention (and its 95% CI).
CI: Confidence interval; MD: Mean Difference; RR: risk ratio
GRADE Working Group grades of evidence :
- High quality: Further research is very unlikely to change our confidence in the estimate of effect - Moderate quality: Further research is likely to have an important impact on our confidence in the estimate of effect and may change the estimate. - Low quality: Further research is very likely to have an important impact on our confidence in the estimate of effect and is likely to change the estimate. - Very low quality: We are very uncertain about the estimate.
RR, risk ratio; CIs, confidence intervals; ROB, risk of bias.

**Table 2.** Summary of uncontrolled studies with aggregated results on the CCP use in immunocompromised patients.

| First author, reference | Population | Diagnosis | COVID-19 | CCP regimen | Days between symptoms and CCP | Previous/concomitant therapies | Outcome | Notes/Conclusions |
| --- | --- | --- | --- | --- | --- | --- | --- | --- |
| Betrains<sup>31</sup> | 5 F, median age 37 y (range 19-67 y) | 5 NHL | Severe COVID-19 | 4 high-titer ($\geq 1:160$ VNT) CCP units | N/A | 1 steroids, 3 HCQ, 1 antibiotics, 2 remdesivir | Increase in nAb titer following CCP<br>Overall mortality rate 1/5 (20%) | Patients with B-cell-depleted lymphomas are ideal candidates for CCP therapy |
| Gharbharan<sup>29</sup> | 25 (15 M, 10 F), median age 53 y (IQR 44-66 y) | 12 NHL<br>1 AL<br>8 AID<br>2 CLL<br>1 MS<br>1 AGG | 19 severe COVID-19,<br>6 mild-moderate COVID-19 | 21 patients 1 CCP unit, 4 patients 2 CCP units | Median 26 d (IQR 15-34 d) | N/A | Overall mortality rate 4/25 (16%) | 24/25 anti-CD20 therapy. The CCP benefit in B-cell depleted patients is present regardless of symptom duration. |
| Greenbaun<sup>30</sup> | 44 (M 25, 19 F), | 17 SC | Severe | 42 patients 1 | N/A | 32 remdesivir, 20 | Overall | Shorter time from COVID- |
| | median age 60<br>(range 37-48 y) | 27 HM | COVID-19 | CCP unit, 2<br>patients 2<br>CCP units | | tocilizumab, 11<br>steroids, 1 anakinra | mortality 12/44<br>(27.3%) | 19 diagnosis to CCP<br>administration ($\leq 3$ days)<br>was associated with better<br>survival |
| Hueso<sup>41</sup> | 17 (12 M, 5 F),<br>median age 58 y<br>(range 35-77 y) | 11 NHL<br>3 CLL<br>1 WM<br>1 CVID<br>1 MS | WHO score 4-7 | CCP units<br>$\geq 1:40$ (VNT)<br>or $> 5.6$<br>(ELISA) | Median 56 d<br>(range 7-83 d) | 8 steroids, 5 HCQ, 4<br>tocilizumab, 3<br>remdesivir, 2 antivirals | Rapid viral<br>clearance<br>following CCP<br>Overall<br>mortality rate<br>1/15 (6.7%) | 15/17 previous treatment<br>with anti-CD20 therapy.<br>CCP is a promising therapy<br>for COVID-19 B-cell<br>depleted patients |
| Jasuja<sup>32</sup> | 22 (N/A) | 22 KTR | N/A | N/A | N/A | HCQ, steroids,<br>antibiotics, remdesivir,<br>tocilizumab | Overall<br>mortality rate<br>7/22 (31.8%) | CCP was associated with no<br>clinical benefit in KTR |
| Jeyaraman<sup>33</sup> | 33 (23 M, 10 F),<br>median age 62 y<br>(range 7-80 y) | 18 NHL<br>4 AL<br>7 MM<br>2 MPD<br>2 MDS | Severe<br>COVID-19 | CCP units<br>$> 1:640$<br>18 patients 1<br>CCP unit, 15<br>patients 2<br>CCP units | Median 4 d<br>(range 2-25 d) | HCQ, steroids,<br>antibiotics, remdesivir,<br>tocilizumab | Overall 42.day<br>mortality rate<br>15/33 (45.5%).<br>No mortality<br>difference<br>between early<br>($< 7$ days) | Study with methodological<br>limitations (retrospective<br>case series) |

|  |  |  |  |  |  |  | versus late CCP<br>transfusion |  |
| --- | --- | --- | --- | --- | --- | --- | --- | --- |
| Levy<sup>34</sup> | 50 (N/A) | 23 NHL<br>9 MM<br>8 CLL<br>1 MPD<br>6 AL<br>2 MDS<br>1 HCL | 40<br>severe/critical<br>COVID-19; 10<br>mild/moderate<br>COVID-19 | N/A | N/A | Steroids, remdesivir | Overall<br>mortality rate<br>16/50 (32%) | CCP was associated with no<br>clinical benefit |
| Ljunquist<sup>35</sup> | 28 (13 M, 15 F),<br>median age 56<br>(range 16-84 y) | 13 HM<br>5 SOT<br>2 PI | Severe<br>COVID-19 | 76 CCP units<br>transfused<br>(median nAb<br>titer 1:141).<br>21 patients<br>received 3<br>CCP units | Median 26 d<br>(range 6-68 d) | 23 steroids, 18<br>remdesivir | Overall 30 days<br>mortality rate<br>6/28 (21.4%) | 50% (14/28) of patients<br>received rituximab.<br>Data not conclusive. |
| Magyari<sup>36</sup> | 20 (13 M, 7 F),<br>median age 56 y<br>(range 27-76 y) | 10 NHL,<br>1 MM<br>4 CLL<br>4 AL | 18 moderate<br>COVID-19<br>(WHO score 4-5) | Median 4<br>CCP units<br>(range 1-15<br>units) | Median 13.5<br>d (range 3-44<br>d) | 17 patients received<br>concomitant<br>remdesivir, 20<br>steroids, 9 antivirals | No COVID-19<br>related deaths<br>were recorded | Anti-CD20 therapy in 13/20<br>patients (65.0%). Clinical<br>benefit of early combined<br>administration of remdesivir |
|  |  | 1 MPD | 2 severe<br>COVID-19<br>(WHO score 8-9) |  |  |  |  | and CCP |
| Tremblay <sup>37</sup> | 24 (14 M, 10 F),<br>median age 69 y<br>(range 31-88 y) | 5 NHL<br>4 MM<br>2 AL<br>1 HL<br>1 MPD<br>1 CLL<br>10 SC | Severe<br>COVID-19 | High titer<br>($\geq 1:320$) CCP<br>units | N/A | 16 HCQ, 15<br>antibiotics, 2<br>remdesivir, 1<br>tocilizumab | Overall<br>mortality rate<br>41.2% (10/24) | Clinical benefit of CCP<br>when administered early in<br>the COVID-19 disease<br>course |
| Rodionov <sup>38</sup> | 14 (7 M, 7 F),<br>median age 65 y<br>(IQR 58-70 y) | 8 SOT<br>4 HSCT<br>2 HM | Median WHO<br>score 5 (range<br>4-6) | Titer $\geq 1:40$<br>CCP units<br>(VNT)<br>1 patient 1<br>CCP unit, 2<br>patients 2<br>CCP units, 11<br>patients 3<br>CCP units | N/A | N/A | Overall<br>mortality 2/14<br>(14.3%).<br>8/14 (57.0%)<br>showed clinical<br>improvement<br>on day 5 after<br>CCP | Immunosuppressed patients<br>are candidates for CCP<br>treatment |
| Sait <sup>39</sup> | 44 (N/A) | SOT | Median WHO<br>score 4 (IQR 3-5) | N/A | N/A | Steroids, remdesivir,<br>antibiotics | Overall<br>mortality 3/44<br>(6.8%) | The use of CC is<br>encouraged in SOT<br>inpatients |
| Weinbergerova<br><sup>40</sup> | 32 (19 M, 13 F),<br>median age 57.7 y<br>(range 25-86 y) | 10 NHL<br>10 AL<br>6 MM<br>3 CLL<br>2 MPD<br>1 AID | 16/32 (50.0%)<br>severe<br>COVID-19 | 2 units high<br>titer ($\geq 1:160$)<br>CCP units | N/A | Steroids, remdesivir | 3/32 (9.4%) | Early COVID-19 treatment<br>with remdesivir + high titer<br>CCP is effective in<br>hematological patients |

**Abbreviations:** AL, acute leukemia; AID, autoimmune disorder; AGG, agammaglobulinemia; CCP, COVID-19 convalescent plasma; CLL, chronic lymphocytic leukemia; CVID, common variable immune deficiency; d, days; ELISA, enzyme-linked immunosorbent assay; F, females; HCL, hairy cell leukemia; HCQ, hydroxychloroquine; HL, Hodgkin's lymphoma; HM, hematological malignancy; HSCT, hematopoietic stem cell transplantation; IQR, interquartile range; KTR, kidney transplant recipients; M, males; MDS, myelodysplastic syndrome; MM, multiple myeloma; MPD, myeloproliferative disorders; MS, multiple sclerosis; N/A, not available; nAb, neutralizing antibody; NHL, non-Hodgkin's lymphoma; PI, primary immunodeficiency; SC, solid cancers; SOT, solid organ transplant; VNT, viral neutralization test; WHO, World Health Organization; WM, Waldenstrom macroglobulinemia; y, years;.

**Figure 2.**
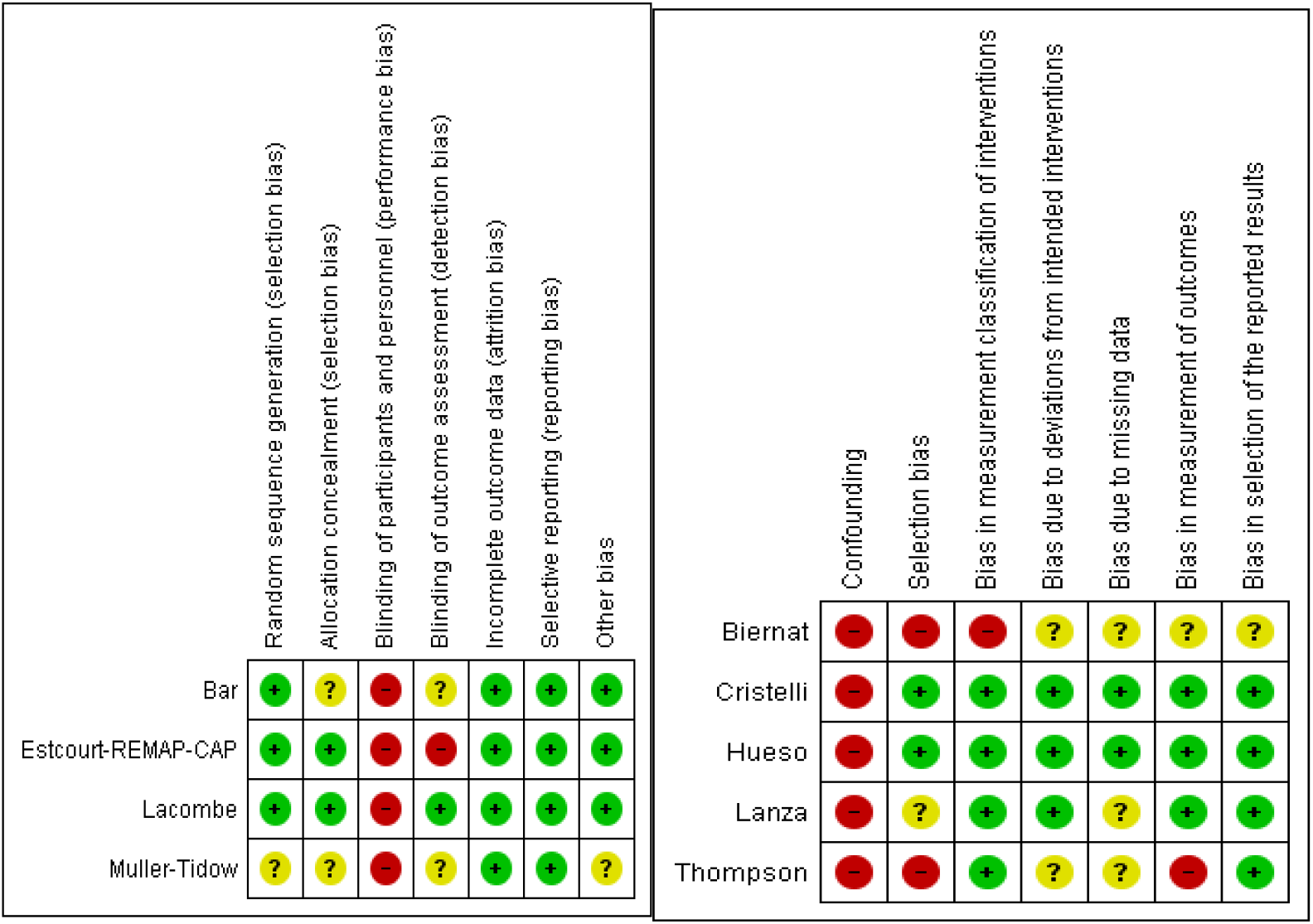
Risk of bias summary for the controlled studies: review authors’ judgements about each risk of bias (ROB) item according to ROBIN-1 tool for each included study. Left: RCTs, analysis according to ROB assessment tool. Right Non-RCTs. Note that although the 3 RCTs were judged at high risk of performance bias because they were open label trials, masking has unclear importance for the outcome mortality, because the risk of ascertainment bias is limited.

**Figure 3.**
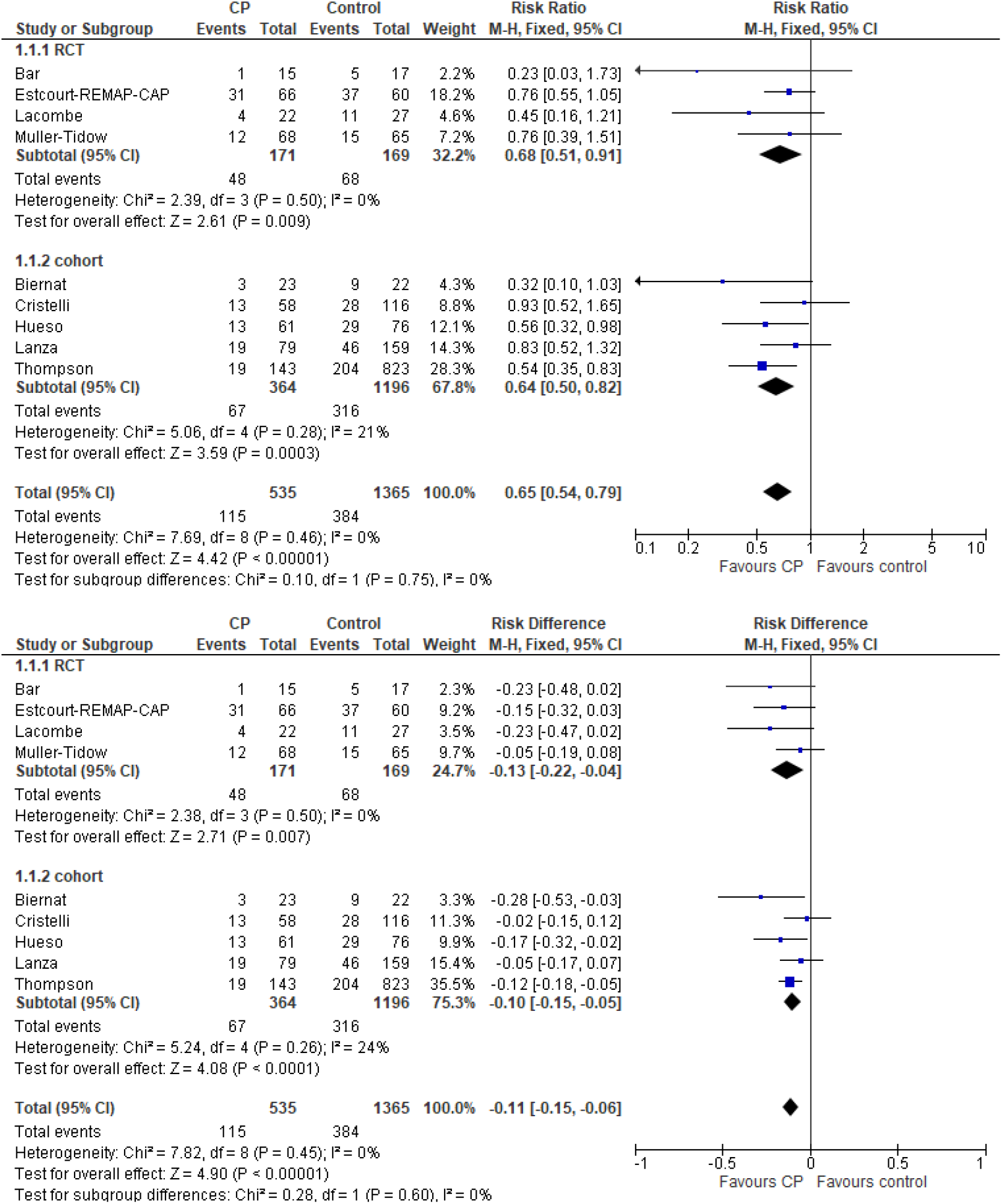
Risk difference (top panel) and risk ratio (bottom panel) for mortality in randomized or cohort-controlled studies included in this systematic review.

The demographic and clinical characteristics of the individual patient data are summarized in Table 3 and are available individually in Supplementary Table 1. The median age was 55 years (range 1-88 years). The male/female ratio was 1.5 (161 males and 105 females). Mean WHO disease severity score was 4.4, with approximately one quarter of patients (51/218) being in ICU on mechanical ventilation. The reported mortality rate was 11.6% (31/265). Forty-seven patients (17.7%) had a primary immunosuppression, mostly agammaglobulinemia (20/47, 42.6%) or common variable immunodeficiency (22/47, 4.8%). The remaining 219 patients had secondary immunosuppression, related to hematological malignancies (134/219, 61.2%), solid cancers (6/219, 2.7%), solid organ transplants (65/219, 29.7%), autoimmune disorders (12/219, 5.5%) or other chronic infectious disease (2/219, 0.9%) (see Table 1). Regarding treatments, the majority of patients (142/265, 53.4%) received steroids as part of the anti-COVID-19 therapy, while 41.7% of them (111/265) received remdesivir and 47% antibiotics (125/265). Approximately one fifth of patients (45/265) also received intravenous immunoglobulin, while a minority of them (11/265, 4.1%) were treated with anti-Spike mAbs: interestingly, in 7 of such cases (7/11, 63.6%), CCP was given as rescue therapy after mAb failure and all 7 patients survived. Nearly 20% of patients (47/265) were under chemotherapy, which in the majority of the cases (35/47, 74.5%) included anti-CD20 mAbs (i.e., rituximab or obinutuzumab), responsible for prolonged B-cell depletion and impaired humoral immune responses. Table 4 summarizes the CCP treatment-related data. The mean number of CCP units transfused per patient was 2.3 (+1.7), while the mean cumulative CCP volume transfused per patient was 460 ml (+372 ml). Unfortunately, it was not possible to calculate the mean nAb titer or to correlate the patients’ outcome with nAb titers due to the wide heterogeneity of tests used (virus neutralization or high-throughput serology). No adverse reactions to CCP were reported. The median time between symptom onset and CCP therapy was 17 days (range 1-132 days), while the median time between hospital admission and CCP therapy was 11 days (0-120 days). The median follow-up period of the patients included in this single patients’ analysis was 19 days (range 4-263 days; data available for 69 patients). Fifty-six percent of the 114 evaluable patients had a rapid (< 5 days) clinical improvement following CCP transfusion. Thirty-one death events were observed (22 male and 9 females). On 126 cases where mortality and total volume were known, 7 death events were found (6 males and 1 female). These data are reported in Table 5 and Figure 4. The 7 death events were observed in the group of 92 patients where the CCP total volume did not exceed 600 ml. The comparison of the mortality when the CCP was under (7 events, 92 patients) or over this level (zero events, 34 patients), however, was not significant. The coefficients of the basic logistic model are reported in Table 6. No inferential significance was obtained. However, the coefficient of “totvol100” had a negative sign, suggesting a decrease of approximately 5% of the starting probability of death for each 100 ml unit added. If the dose-response effect was real, with 600 ml of CCP the mortality reduction would be 26.64%, with 1200 ml would be 46.48%, and with 1800 ml would be 61.12%. Of course, this would need confirmation by a more extended number of observations. Adding to the basic model the above listed covariates, “Age”, “Mechanical Ventilation”, “Category=autoimmune”, “Category=solid cancer”, and “Anti-Spike mAbs” appeared to predict a significant increase of mortality. By logistic model, the evaluation of the role of anti-CD20 was impossible since anti-CD20 = “yes” predicted failure (survival) perfectly (nobody died if treated with anti-CD20 if CCP total volume information was available). This happened with “Rapid improvement”, “(Hydroxy)chloroquine”, “Category=common”, “Category=humoral” as well. In contrast, failure to receive steroids or antibiotics predicted survival perfectly; in other words, deaths were found only in treated people.

**Table 3.**
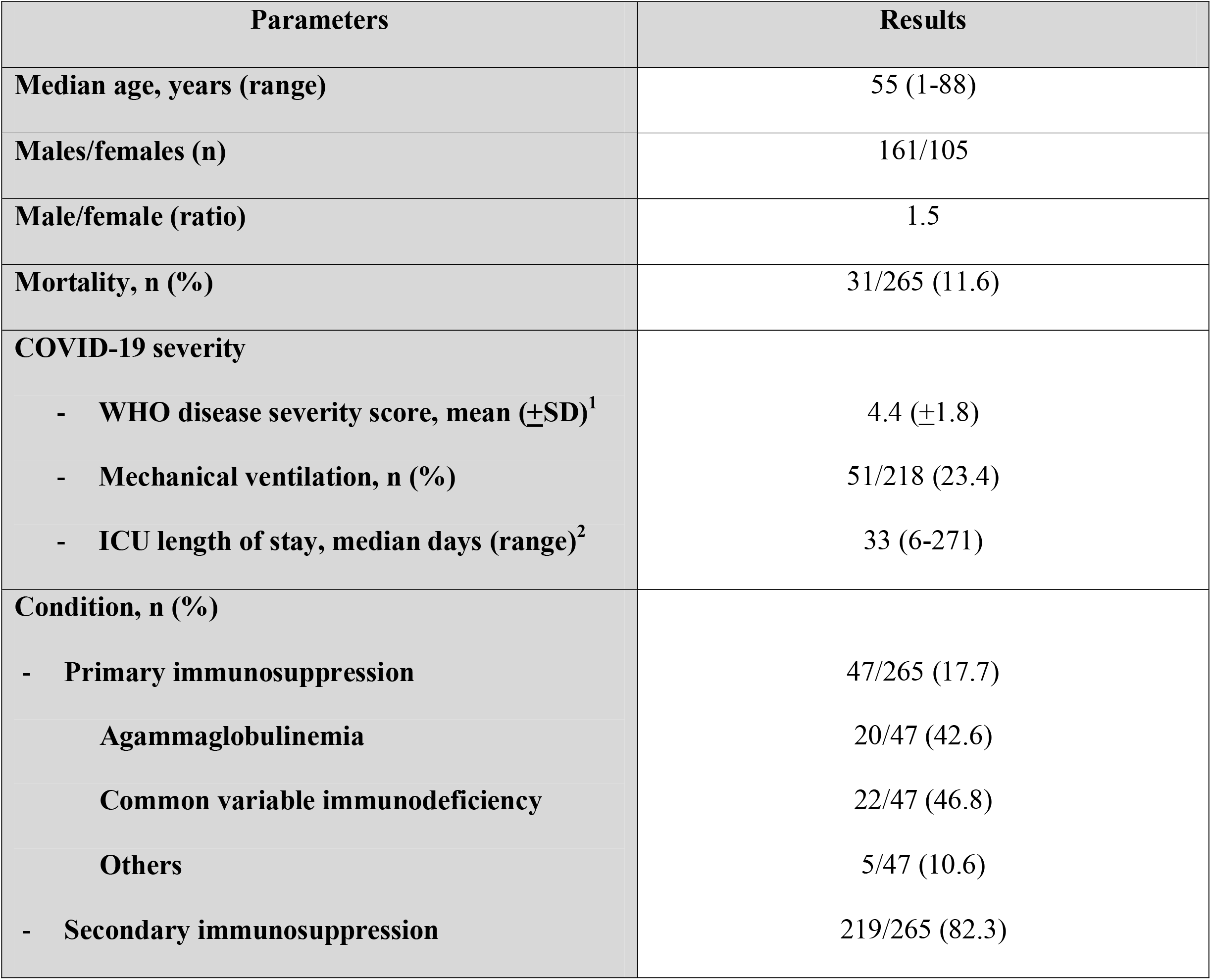
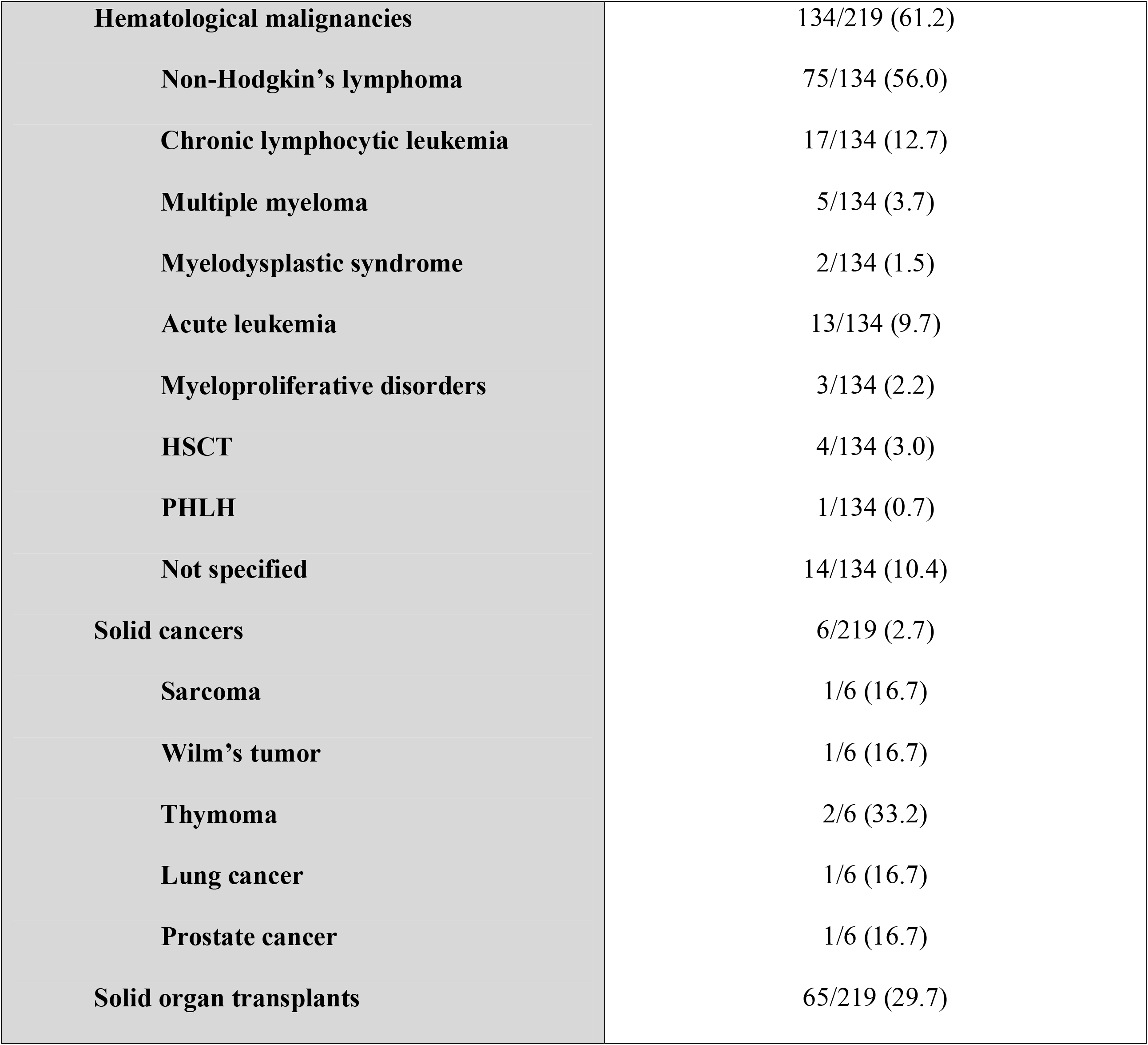
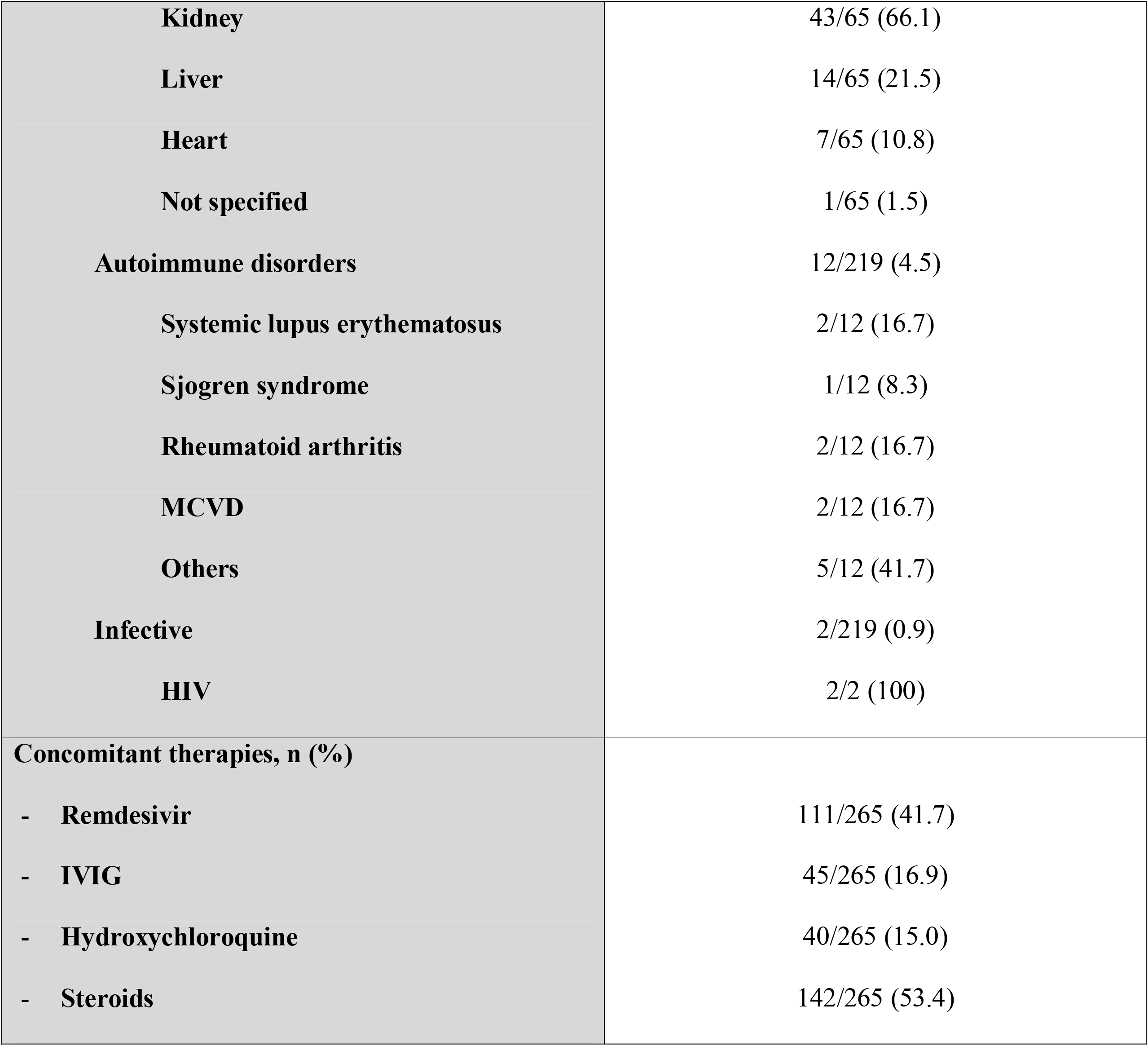
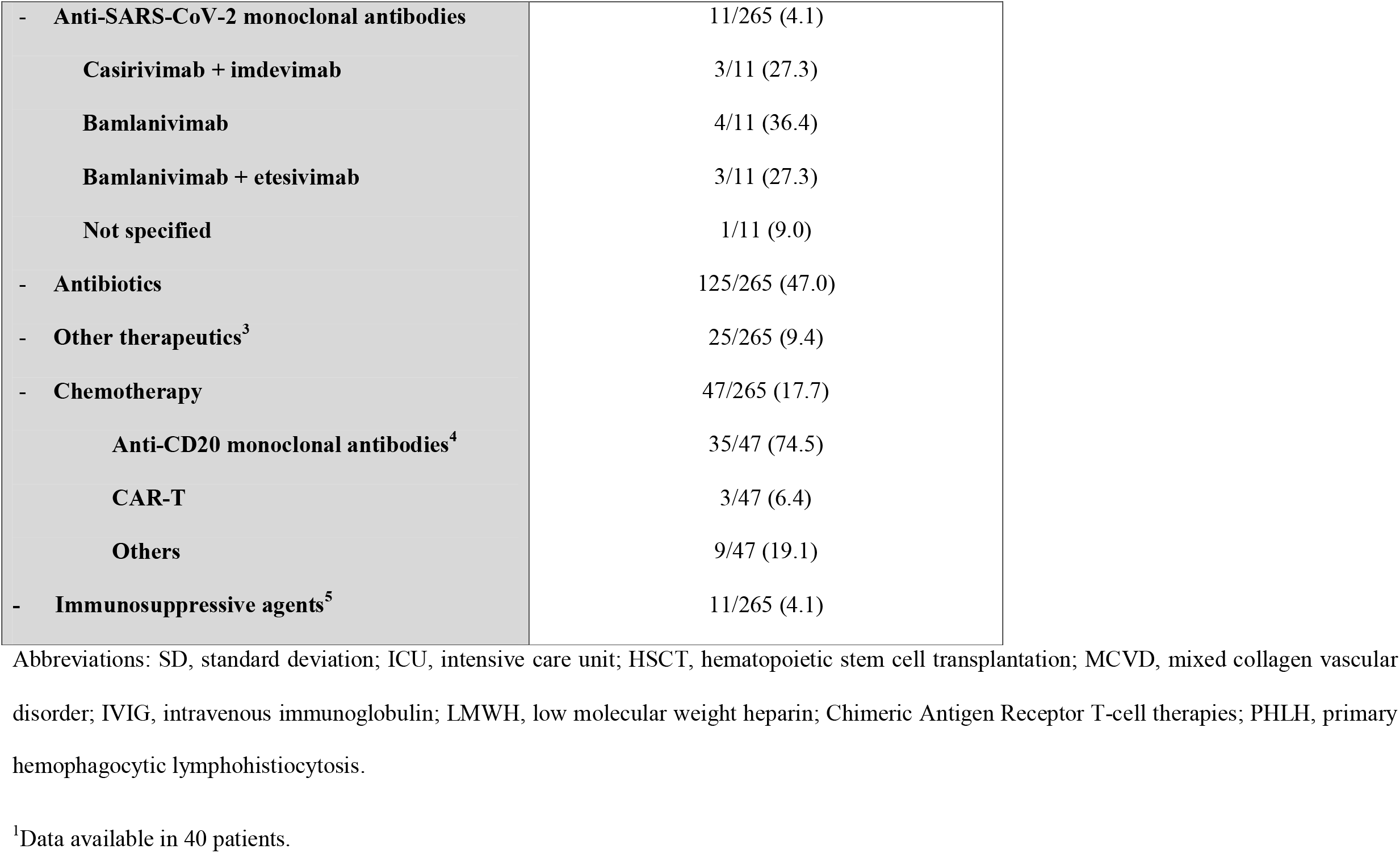
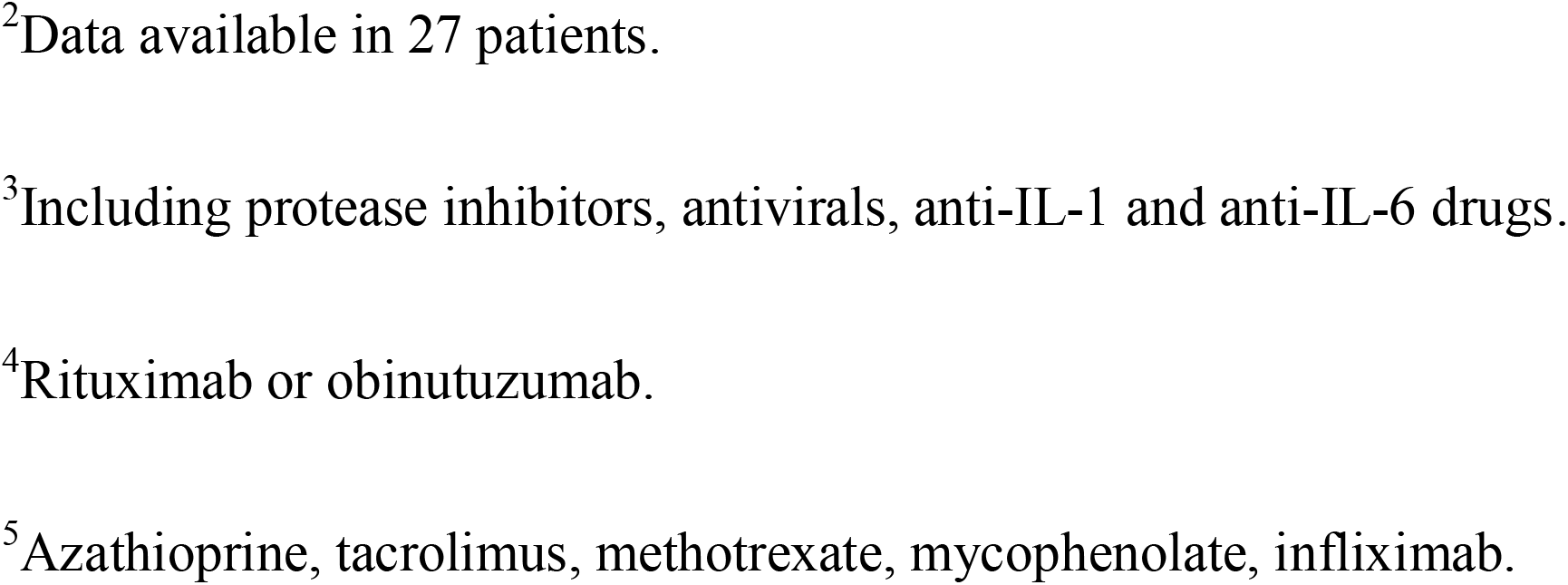
Demographic and clinical characteristics of the 265 patients included in the individual patients’ analysis.

| Parameters | Results |
| --- | --- |
| Median age, years (range) | 55 (1-88) |
| Males/females (n) | 161/105 |
| Male/female (ratio) | 1.5 |
| Mortality, n (%) | 31/265 (11.6) |
| COVID-19 severity |  |
| - WHO disease severity score, mean (±SD)<sup>1</sup> | 4.4 (±1.8) |
| - Mechanical ventilation, n (%) | 51/218 (23.4) |
| - ICU length of stay, median days (range)<sup>2</sup> | 33 (6-271) |
| Condition, n (%) |  |
| - Primary immunosuppression | 47/265 (17.7) |
| Agammaglobulinemia | 20/47 (42.6) |
| Common variable immunodeficiency | 22/47 (46.8) |
| Others | 5/47 (10.6) |
| - Secondary immunosuppression | 219/265 (82.3) |
| **Hematological malignancies** | 134/219 (61.2) |
| **Non-Hodgkin's lymphoma** | 75/134 (56.0) |
| **Chronic lymphocytic leukemia** | 17/134 (12.7) |
| **Multiple myeloma** | 5/134 (3.7) |
| **Myelodysplastic syndrome** | 2/134 (1.5) |
| **Acute leukemia** | 13/134 (9.7) |
| **Myeloproliferative disorders** | 3/134 (2.2) |
| **HSCT** | 4/134 (3.0) |
| **PHLH** | 1/134 (0.7) |
| **Not specified** | 14/134 (10.4) |
| **Solid cancers** | 6/219 (2.7) |
| **Sarcoma** | 1/6 (16.7) |
| **Wilm's tumor** | 1/6 (16.7) |
| **Thymoma** | 2/6 (33.2) |
| **Lung cancer** | 1/6 (16.7) |
| **Prostate cancer** | 1/6 (16.7) |
| **Solid organ transplants** | 65/219 (29.7) |
| **Kidney** | 43/65 (66.1) |
| **Liver** | 14/65 (21.5) |
| **Heart** | 7/65 (10.8) |
| **Not specified** | 1/65 (1.5) |
| **Autoimmune disorders** | 12/219 (4.5) |
| **Systemic lupus erythematosus** | 2/12 (16.7) |
| **Sjogren syndrome** | 1/12 (8.3) |
| **Rheumatoid arthritis** | 2/12 (16.7) |
| **MCVD** | 2/12 (16.7) |
| **Others** | 5/12 (41.7) |
| **Infective** | 2/219 (0.9) |
| **HIV** | 2/2 (100) |
| **Concomitant therapies, n (%)** |  |
| - **Remdesivir** | 111/265 (41.7) |
| - **IVIg** | 45/265 (16.9) |
| - **Hydroxychloroquine** | 40/265 (15.0) |
| - **Steroids** | 142/265 (53.4) |
| - **Anti-SARS-CoV-2 monoclonal antibodies** | 11/265 (4.1) |
| **Casirivimab + imdevimab** | 3/11 (27.3) |
| **Bamlanivimab** | 4/11 (36.4) |
| **Bamlanivimab + etesivimab** | 3/11 (27.3) |
| **Not specified** | 1/11 (9.0) |
| - **Antibiotics** | 125/265 (47.0) |
| - **Other therapeutics<sup>3</sup>** | 25/265 (9.4) |
| - **Chemotherapy** | 47/265 (17.7) |
| **Anti-CD20 monoclonal antibodies<sup>4</sup>** | 35/47 (74.5) |
| **CAR-T** | 3/47 (6.4) |
| **Others** | 9/47 (19.1) |
| - **Immunosuppressive agents<sup>5</sup>** | 11/265 (4.1) |

Abbreviations: SD, standard deviation; ICU, intensive care unit; HSCT, hematopoietic stem cell transplantation; MCVD, mixed collagen vascular disorder; IVIG, intravenous immunoglobulin; LMWH, low molecular weight heparin; Chimeric Antigen Receptor T-cell therapies; PHLH, primary hemophagocytic lymphohistiocytosis.
<sup>1</sup>Data available in 40 patients.
<sup>2</sup>Data available in 27 patients.
<sup>3</sup>Including protease inhibitors, antivirals, anti-IL-1 and anti-IL-6 drugs.
<sup>4</sup>Rituximab or obinutuzumab.
<sup>5</sup>Azathioprine, tacrolimus, methotrexate, mycophenolate, infliximab.

**Table 4.** COVID-19 convalescent plasma treatment-related data in 265 individual patients.

| Parameters | Information<br>available for no.<br>patients | Results |
| --- | --- | --- |
| CCP transfused units, mean (±SD; range)<sup>1</sup> | 153 | 2.3 (±1.7; 1-11) |
| CCP total transfused volume (ml)<sup>2</sup> | 126 | 460 (±372.0; 200-1800) |
| Days between symptom onset and CCP therapy,<br>median (range)<sup>3</sup> | 188 | 17 (1-132) |
| Days between hospital admission and CCP therapy, median (range)<sup>4</sup> | 101 | 11 (0-120) |
| Post-CCP rapid improvement in oxygen supplementation (≤ 5 days), n<br>(%) | 114 | 64/114 (56.1) |

**Table 5.** Mortality under different levels of CCP treatment, as total volume from <= 200 ml to >1800 ml. Seven death events (5.56%) were observed on 125 patients with available CCP total volume information.

| Total Volume | Mortality | Patients |
| --- | --- | --- |
| $\leq 200$ | 0 | 19 |
| $>200-400$ | 3 | 43 |
| $>400-600$ | 4 | 29 |
| $>600-800$ | 0 | 12 |
| $>800-1000$ | 0 | 12 |
| $>1000-1200$ | 0 | 1 |
| $>1200-1400$ | 0 | 0 |
| $>1400-1600$ | 0 | 4 |
| $>1600-1800$ | 0 | 4 |
| $>1800$ | 0 | 1 |
| Total | 7 | 125 |

**Table 6.** Logistic regression, table of coefficients. Mortality was the dependent variable, CCP total volume was the predictor. The CCP total volume was expressed in units of 100 ml (“totvol100”). Number of obs = 125, log likelihood = -26.896492, LR chi2(1) = 0.28, P = 0.5994, Pseudo R2 = 0.0051.

| Mortality | Coefficient | Std. err. | z | P | 95% conf. interval |  |
| --- | --- | --- | --- | --- | --- | --- |
| totvol100 | -0.0551 | 0.1156 | -0.48 | 0.634 | -0.2818 | 0.1716 |
| _cons | -2.5370 | 0.6998 | -3.63 | 0.000 | -3.9086 | -1.1654 |

**Figure 4.**
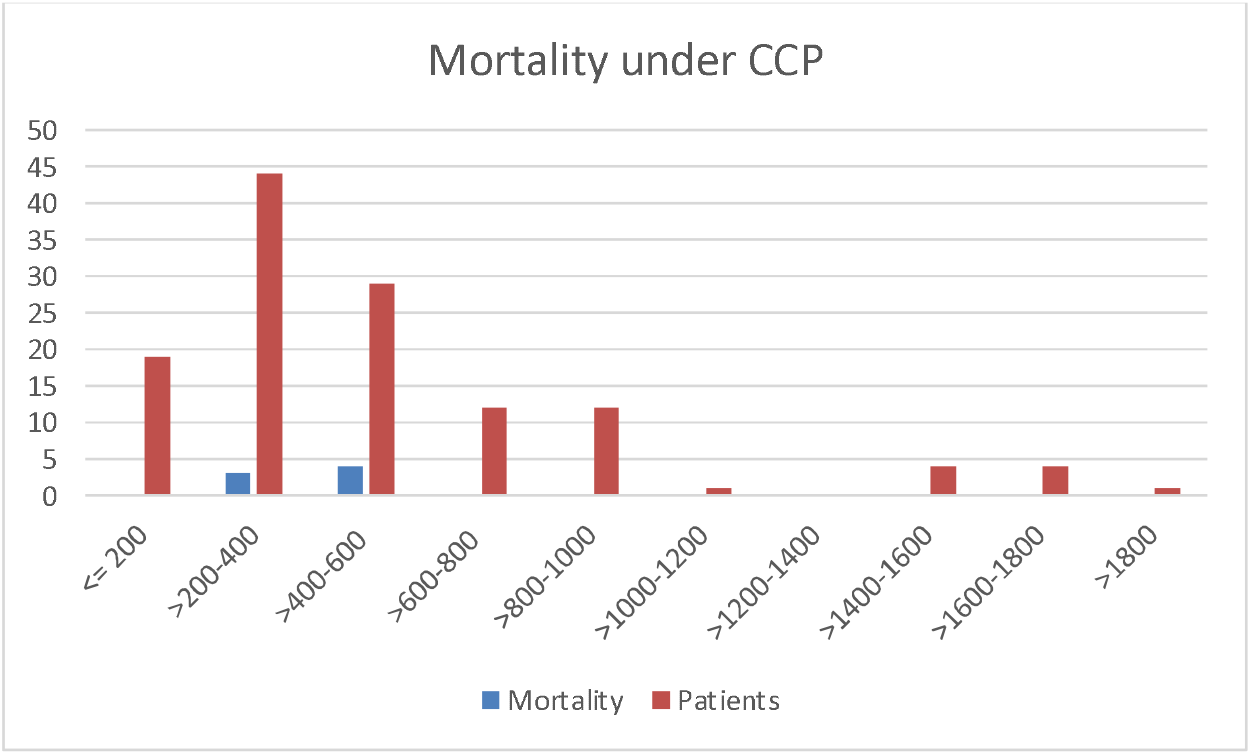
Mortality under different levels of CCP treatment in 265 individual patients, as total volume from <= 200 ml to >1800 ml. Blue bars: death incidence. Orange bars: number of patients at risk at each level of CCP.

**Figure 5.**
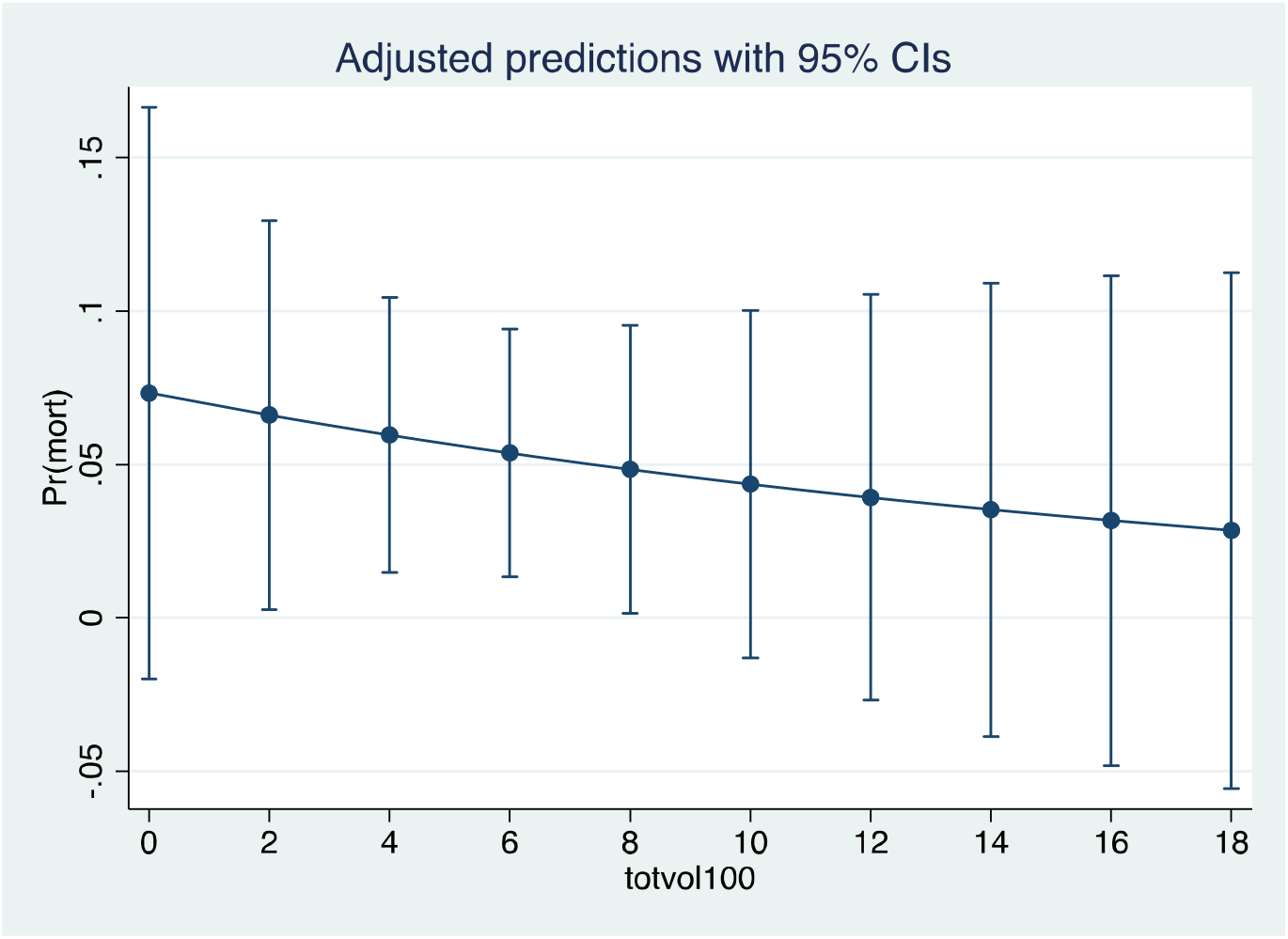
Predicted probability of death at various levels of CCP total volume, after logistic regression, basic model. The CCP total volume (“totvol100”) is expressed as 100 ml units. The large confidence intervals pinpoint the need for a confirmation by a more extended number of observations.

The total sample size calculated by power analysis for RR = 0.75 was 4,024, for RR = 0.5 was 870.

## Discussion

Several scientific societies (e.g., ECIL-9 ^166^, CDC/IDSA^167^ and AABB ^168^ have recently revised their guidelines to recommend usage of CCP in immunocompromised patients, expecially after Omicron sublineages progressively defeated mAb therapies authorized so far.

The hypothesis of a significant beneficial effect of CCP on mortality in immunocompromised patients cannot be definitively demonstrated with the present data, but very strong elements support its efficacy. The efficacy of antibody-based therapies for immunocompetent individuals is predicated on early administration with sufficient dosage ^169^. This principle was validated by the experience of CCP in COVID-19 ^9^. While several immunocompromised cases have been treated with CCP derivatives (hyperimmune immunoglobulins) ^170^, CCP is superior in turnaround times and inclusion of classes other than IgG ^171^. However, we note that the immunosuppressed patients in this study were treated relatively late after the initial symptoms (17 d) and hospital admission (11 d) and yet our analysis suggests a benefit for CCP. For life-threatening COVID-19 the pathogenesis involves exuberant tissue-damaging inflammatory responses that follow an initial viral phase. Antibody-based therapies function primarily as antiviral agents and are much less likely to be affected in individuals who are in the inflammatory phase. However, immunocompromised individuals are generally unable to mount strong antibody or inflammatory responses and often cannot clear SARS-CoV-2. Hence, immunosuppressed patients represent a biologically different population from the immunocompetent population where antibody-based therapies may retain efficacy late into the course of disease.

The efficacy of CCP in immunosuppressed patients that had reported symptoms for weeks or months paves the way to the hypothesis that CCP retains clinical efficacy until the recipient is seronegative and there is no irreversible parenchymal damage. The recently reopened CCP arm of the REMAP-CAP randomized controlled trial in UK will specifically target immunocompromised patients in ICU focusing on CCP from vaccinated donors (so-called VaxCCP or “hybrid” plasma) ^172^. While most studies reported in this systematic review used CCP from unvaccinated donors (with a few exceptions ^125,152^), it is noteworthy that VaxCCP is nowadays widely available from regular donors, retains higher nAb titers and efficacy against most Omicron sublineages than standard CCP ^173^.

## Data Availability

All data produced in the present study are available upon reasonable request to the authors

## Data availability statement

This secondary research did not generate original data, which remain available at the cited references.

## Abbreviations

CCP: COVID-19 convalescent plasma;
nAb: neutralizing antibodies;
VOC: variant of concern.

## Acknowledgements

We are grateful to Dr.Thomas Hueso who contributed to data analysis.

